# Associations of alcohol use in early and middle adulthood with mid- and late-life cognition - a synthetic cohort approach

**DOI:** 10.64898/2026.02.27.26346914

**Authors:** Peter T Buto, Scott C Zimmerman, Katrina Kezios, Adina Zeki Al Hazzouri, M Maria Glymour

## Abstract

**INTRODUCTION:** Early adulthood alcohol use may influence later life memory, but few studies assessed this association. Synthesizing two cohorts, we estimated effects of prospectively reported early life alcohol use on memory across middle and later adulthood.

**METHODS:** We used National Longitudinal Study of Youth 1979 (NLSY79, n=12221, alcohol reports ages 18-26), Health and Retirement Study (HRS, age 50-56 at enrollment, n=21,263), and a synthetic cohort matching early life alcohol use among NLSY79 participants to later life memory among HRS participants. Covariate-adjusted linear mixed models regressed memory (word list recall) on alcohol use category (none, light/moderate, heavy).

**RESULTS:** Early adulthood alcohol use category was not associated with midlife memory in the NLSY79 cohort or with middle to late life memory or memory change in the synthetic cohort.

**DISCUSSION:** Early adulthood alcohol use has little association with memory function at midlife or later.

## Introduction

Alcohol use is a common, modifiable, and potentially impactful risk factor for dementia. The Lancet’s Lancet Commission on Dementia identified that up to 1% of dementia cases in the United States may be prevented by eliminating excessive alcohol use.^1^ Extant evidence however, does not clearly delineate whether the effects of alcohol consumption vary by lifecourse timing or whether major effects are limited to heavy alcohol consumption.^2,3^ Drinking patterns across the life course may have different effects on cognitive aging: critical periods of alcohol use, particularly during late adolescence and young adulthood, have been identified for other health outcomes.^2,4^ Few data sources include both long-term information on alcohol use and longitudinal cognitive aging data.^5^

The National Longitudinal Study of Youth 1979 (NLSY79) prospectively recorded alcohol consumption from late adolescence or early adulthood through middle age but includes only a maximum of two memory assessments in middle age.^6^ In contrast, the Health and Retirement Study (HRS) has decades of longitudinal memory information, but cohort eligibility begins at age 50, so no information on early adulthood alcohol consumption is available.^7^

Recent methods in lifecourse epidemiology emphasize the potential for synthetic cohorts to address such data gaps.^8^ To examine the effects of adolescent alcohol use on midlife memory score and mid-to-late life memory decline, we used NLSY79, HRS, and six synthetic cohorts combining the NLSY79 and HRS. Synthetic cohort construction uses an early-life cohort (NLSY79, in which the exposure is measured) to stand in for the unobserved earlier-life drinking histories of similar participants in a late-life cohort (HRS, in which the outcome is measured).

We first investigated the association between alcohol use and memory score within the available age ranges of each cohort separately. In NLSY79 alone, we estimated associations between early adulthood alcohol use at eight waves with midlife memory score and memory decline between two time points. In HRS alone, we estimated associations of midlife alcohol use with midlife memory score and mid-to-late life memory decline over up to 12 survey waves. In the synthetic cohort, we then estimated the association between alcohol use at the same eight early adulthood time points from NLSY79 with midlife memory and mid-to-late life memory decline, as measured in HRS.

## Methods

### Samples

The NLSY79 is a closed cohort study of 12,686 participants ages 14-22 when enrolled in 1979. Over the course of follow-up, data on behaviors, occupation, education, socio-economic status, etc. were collected prospectively to study the life-course experience.^6^

The HRS is an open cohort that periodically recruits new participants to maintain a probability sample of community-residing individuals over age 50. Participants complete biennial interviews on social circumstances and health, including memory score.^7^ Cognitive testing procedures differed in early HRS waves compared to later waves and the sample was not nationally representative of all relevant birth cohorts. We therefore used data from participants who joined HRS in the 1998 wave or later.

### Alcohol

We used data from (n=7,540) NLSY79 participants with alcohol information reported at 8 time points from 1983 (ages 18-26) through 2006 (ages 41-48), to classify alcohol use from early adulthood to mid-life into sex-specific categories, based on estimated weekly alcohol consumption. To allow for harmonized alcohol categories across waves, participants were classified as nondrinkers, light-to-moderate drinkers (no more than 7 drinks per week for women and no more than 14 drinks per week for men, without reported heavy episodic drinking defined as 6 or more drinks any point in the last 30 days, regardless of gender), or heavy drinkers (more than 7 drinks per week for women or 14 drinks for men, or any reported heavy episodic drinking). Due to differences in fielded questions, these alcohol measures varied slightly across waves: from 1988 onward we calculated days drank/week from drinks/month.

Current midlife alcohol consumption in HRS was examined at each participant’s cohort entry wave between 1998-2020 (n=21,263). HRS participants reported alcohol use in days drank per week over the past 3 months and drinks consumed per day on those days. We used these data to create an alcohol measure harmonized with those described in the NLSY79-Alcohol section. While this measure is similar to the Substance Abuse and Mental Health Services Administration (SAMHSA) drinking levels classification,^9,10^ it was not possible to include gender-specific thresholds for heavy episodic drinking because the HRS questions did not have enough resolution.

### Memory data

The NLSY79 participants were administered their first cognitive battery in the year they turned 48. This battery included a 10-word list immediate recall task and a delayed recall task of the same list. We summarized midlife memory score as the sum of immediate and delayed word recall scores. Each recall score was coded as the number of correctly recalled words out of a list of ten. We created a summed recall score variable by adding scores for immediate and delayed recall. A second cognitive battery was administered in 2020, with an average follow-up time of 10 years between the first and second cognitive assessment.

HRS participants were also administered an immediate and delayed word recall test using a different list of 10 words.^11^ We summarized memory scores in HRS following the rules for NLSY79. We used memory scores from 1998-2020 in HRS. In analyses conducted using HRS data alone, the available sample size with information on midlife alcohol use and at least one subsequent memory assessment was 13,090 participants.

We pre-processed memory scores to account for repeated testing following the approach used in Chen et al. 2023, regressing memory at each wave on indicators for the number of previous assessments (0, 1, 2 or more) and using the coefficients from these regressions to back out the effects from the first and second exposure to the cognitive battery.^12^ This outcome can be interpreted as the memory score after participants have become accustomed to the cognitive testing procedure.

### Matching to create the synthetic cohorts

We created one synthetic cohorts for each exposure-year in NLSY79 (six synthetic cohorts) by matching up to ten NLSY79 participants to a given HRS participant based on a fixed set of time-invariant confounders as well as a varying set of time-varying mediators hypothesized to capture pathways linking alcohol consumption at each time period (as reported in NLSY79) with mid-to-late life memory (as reported in HRS). In other words, we selected matching variables hypothesized to be important confounders or mediators of the relationship.

#### Matching confounders

For all synthetic cohorts, each HRS participant was exact-matched to all NLSY79 participants who shared the same confounders (gender, race/ethnicity, knowledge of parental education, and US nativity). To ensure participants were matched at similar ages and time periods within each of the synthetic cohorts defined by exposure-year in NLSY79, we also matched on birth year [within a window of 5 years] as well as age [within a window of 5 years] at the time period in which the NLSY79 synthetic cohort and HRS cohort began to have age-overlap (Table 1).

**Table 1:** Sample characteristics for NLSY and HRS prior to matching.

|  | Descriptive Statistic (mean (sd) or n(%)) |  |  |  |  |  |  |
| --- | --- | --- | --- | --- | --- | --- | --- |
|  | NLSY |  |  |  |  |  | HRS |
| Variable | 1983 | 1985 | 1988 | 1994 | 2002 | 2006 |  |
| Sample Size | 12221 | 10894 | 10465 | 8889 | 7723 | 7653 | 21263 |
| Age at Interview | 22.3<br>(2.3) | 24.1<br>(2.2) | 27.6<br>(2.2) | 33.5<br>(2.2) | 41.4<br>(2.2) | 45.2<br>(2.2) | 55.4<br>(7.9) |
| Birth Year | 1960.4<br>(2.2) | 1960.6<br>(2.2) | 1960.6<br>(2.2) | 1960.6<br>(2.2) | 1960.6<br>(2.2) | 1960.6<br>(2.2) | 1951.9<br>(12.0) |
| Alcohol Binge | 0.6<br>(0.8) | 0.3<br>(0.7) | 0.5<br>(0.8) | 0.4<br>(0.8) | 0.2<br>(0.6) | 0.2<br>(0.7) | 0.3<br>(0.7) |
| Immediate Recall Score | Not<br>Measur<br>ed | Not<br>Measur<br>ed | Not<br>Measur<br>ed | Not<br>Measur<br>ed | Not<br>Measur<br>ed | 5.9<br>(1.8) | 5.7<br>(1.6) |
| Delayed Recall Score | Not<br>Measur<br>ed | Not<br>Measur<br>ed | Not<br>Measur<br>ed | Not<br>Measur<br>ed | Not<br>Measur<br>ed | 4.8<br>(2.0) | 4.6<br>(1.9) |
| Alcohol: Drinks Per Month | Not<br>Measur<br>ed | Not<br>Measur<br>ed | 25.6<br>(45.1) | 23.3<br>(45.1) | 21.8<br>(38.0) | 20.7<br>(38.2) | 13.5<br>(33.4) |
| Alcohol: Ever | 11424<br>(93.5%) | 10298<br>(94.5%) | 9974<br>(95.3%) | 7217<br>(81.2%) | 6302<br>(81.6%) | 6223<br>(81.3%) | 18449<br>(86.8%) |
| Alcohol: Current | 8069<br>(66%) | 7215<br>(66.2%) | 6996<br>(66.9%) | 5319<br>(59.8%) | 4150<br>(53.7%) | 3953<br>(51.7%) | 14093<br>(66.3%) |
| BMI (kg/m<sup>2</sup>) | 23.0<br>(3.7) | 23.8<br>(4.2) | 24.8<br>(4.7) | 26.6<br>(5.5) | 28.3<br>(6.1) | 28.9<br>(6.1) | 29.1<br>(6.5) |
| Alcohol: Do you ever drink first thing in the morning? | 0 (0%) | 185<br>(1.7%) | 186<br>(1.8%) | 158<br>(1.8%) | 138<br>(1.8%) | 145<br>(1.9%) | 1330<br>(6.3%) |
| Diagnosis: Cancer | Not<br>Measur<br>ed | Not<br>Measur<br>ed | Not<br>Measur<br>ed | Not<br>Measur<br>ed | 100<br>(1.3%) | 155<br>(2%) | 1525<br>(7.2%) |
| CES-D (modified 6pt scale) | Not<br>Measur | Not<br>Measur | Not<br>Measur | Not<br>Measur | 1.7<br>(1.9) | 1.7<br>(1.9) | 1.4<br>(1.7) |
|  | ed | ed | ed | ed |  |  |  |
| Father's Highest Education | 10.9<br>(3.8) | 10.8<br>(3.8) | 10.8<br>(3.8) | 10.8<br>(3.8) | 10.8<br>(3.8) | 10.8<br>(3.8) | 10.1<br>(4.4) |
| Diagnosis: Diabetes | Not Measured | Not Measured | Not Measured | Not Measured | 283<br>(3.7%) | 410<br>(5.4%) | 3499<br>(16.5%) |
| Education (Categorical) | 1.7<br>(0.7) | 1.8<br>(0.7) | 1.9<br>(0.7) | 1.9<br>(0.7) | 1.9<br>(0.7) | 1.9<br>(0.7) | 2.0<br>(0.8) |
| General Health (Categorical) | Not Measured | Not Measured | Not Measured | Not Measured | 1.3<br>(1.0) | 1.4<br>(1.0) | 1.8<br>(1.2) |
| History of a Heart Problem | Not Measured | Not Measured | Not Measured | Not Measured | 146<br>(1.9%) | 212<br>(2.8%) | 2735<br>(12.9%) |
| Height (Inches) | 67.1<br>(4.1) | 67.1<br>(4.1) | 67.1<br>(4.1) | 67.1<br>(4.1) | 67.0<br>(4.1) | 67.2<br>(4.1) | 66.5<br>(4.1) |
| History of Hypertension | Not Measured | Not Measured | Not Measured | Not Measured | 901<br>(11.7%) | 1321<br>(17.3%) | 9181<br>(43.2%) |
| Income Per Person (Log10) | 1.2<br>(1.2) | 1.4<br>(1.3) | 1.6<br>(1.4) | 1.5<br>(1.3) | 1.6<br>(1.3) | 1.7<br>(1.4) | 2.1<br>(1.1) |
| Light Physical Activity | Not Measured | Not Measured | Not Measured | Not Measured | 5605<br>(72.6%) | 5475<br>(71.5%) | 11721<br>(55.1%) |
| Marital Status: Married | 4386<br>(35.9%) | 4740<br>(43.5%) | 5832<br>(55.7%) | 5618<br>(63.2%) | 4530<br>(58.7%) | 4374<br>(57.2%) | 3505<br>(16.5%) |
| Marital Status: Never | 7241<br>(59.3%) | 5402<br>(49.6%) | 3521<br>(33.6%) | 1923<br>(21.6%) | 1439<br>(18.6%) | 1351<br>(17.7%) | 15205<br>(71.5%) |
| Marital Status: Separated, Divorced, Absent | 582<br>(4.8%) | 739<br>(6.8%) | 1085<br>(10.4%) | 1307<br>(14.7%) | 1682<br>(21.8%) | 1820<br>(23.8%) | 1460<br>(6.9%) |
| Marital Status: widowed | 12<br>(0.1%) | 13<br>(0.1%) | 27<br>(0.3%) | 41<br>(0.5%) | 72<br>(0.9%) | 108<br>(1.4%) | 1072<br>(5%) |
| Military Service/Veteran Status | 2071<br>(16.9%) | 1166<br>(10.7%) | 1157<br>(11.1%) | 1065<br>(12%) | 915<br>(11.8%) | 919<br>(12%) | 3164<br>(14.9%) |
| Mother's Highest Education | 10.9<br>(3.1) | 10.8<br>(3.2) | 10.8<br>(3.2) | 10.8<br>(3.2) | 10.8<br>(3.2) | 10.8<br>(3.2) | 10.3<br>(4.1) |
| Self-rated cognition | Not Measured | Not Measured | Not Measured | Not Measured | Not Measured | 2.6<br>(1.0) | 2.6<br>(1.0) |
| Self-rated cognition compared to past | Not Measured | Not Measured | Not Measured | Not Measured | Not Measured | 2.1<br>(0.5) | 2.1<br>(0.5) |
| Religion: Catholic | 3796<br>(31.1%) | 3429<br>(31.5%) | 3254<br>(31.1%) | 2910<br>(32.7%) | 2035<br>(26.3%) | 2037<br>(26.6%) | 5602<br>(26.3%) |
| Religion: Jewish | 97<br>(0.8%) | 87<br>(0.8%) | 86<br>(0.8%) | 64<br>(0.7%) | 54<br>(0.7%) | 52<br>(0.7%) | 286<br>(1.3%) |
| Religion: None | 1422<br>(11.6%) | 1235<br>(11.3%) | 1190<br>(11.4%) | 954<br>(10.7%) | 827<br>(10.7%) | 823<br>(10.8%) | 739<br>(3.5%) |
| Religion: Other | 221<br>(1.8%) | 202<br>(1.9%) | 191<br>(1.8%) | 147<br>(1.7%) | 606<br>(7.8%) | 584<br>(7.6%) | 11703<br>(55%) |
| Religion: Protestant | 6683<br>(54.7%) | 5939<br>(54.5%) | 5742<br>(54.9%) | 4812<br>(54.1%) | 4201<br>(54.4%) | 4157<br>(54.3%) | 2859<br>(13.4%) |
| Serial 7s | Not Measured | Not Measured | Not Measured | Not Measured | Not Measured | 3.7<br>(1.5) | 3.6<br>(1.5) |
| Ever Smoked | Not Measured | Not Measured | Not Measured | Not Measured | Not Measured | Not Measured | 11853<br>(55.7%) |
| Current Smoke | Not Measured | Not Measured | Not Measured | Not Measured | Not Measured | Not Measured | 4936<br>(23.2%) |
| Physical Activity: Vigorous | Not Measured | Not Measured | Not Measured | Not Measured | 2882<br>(37.3%) | 2809<br>(36.7%) | 7165<br>(33.7%) |
| Weight (lbs) | 147.7<br>(30.3) | 152.8<br>(32.9) | 159.6<br>(35.9) | 170.8<br>(39.6) | 181.5<br>(43.0) | 185.9<br>(44.3) | 183.5<br>(44.5) |

Summary NLSY descriptive statistics are from each exposure round of NLSY. Summary HRS descriptive statistics are from the first wave with memory measured for each participant from HRS Years 1998-2020

#### Matching mediators

We further matched HRS participants to NLSY79 participants based on a set of midlife mediators hypothesized to link earlier alcohol use with later memory outcomes. We considered the following midlife mediators as potential matching variables: mid-life alcohol use, physical activity (moderate and vigorous separately), education, marital status, history of any military service, current religion, depression, history of health conditions (diabetes, hypertension, cancer, & heart problems), smoking history at each wave. Where necessary, we determined appropriate common codings and created harmonized versions of variables.

In order to maximize statistical power, we prioritized matching on the minimal set of variables most strongly associated with both the exposure and the outcome. To do this, we first ran unadjusted linear regressions between each matching mediators with the exposure and the outcome. We identified the matching variable with the largest R^2^ and then included that variable in the adjustment set for another round of regressions in the remaining matching variables.

Having ranked the matching mediators, we then determined the optimal matching set. This entails balancing the validity advantages of a comprehensive list against the power cost of additional matching variables. We seek to find the smallest matching set with a bias no larger than that of the estimate as defined by a dataset imputed with all of the mediating variables. For example, for the association between alcohol consumption in 1988 with mid-to-late life cognition, in addition to all time-invariant matching confounders above, the optimal matching set among all candidate mediators matched on midlife alcohol consumption, immediate word recall score, and delayed word recall score.

We used exact matching for categorical variables. For continuous variables, we quantified the similarity between the HRS participant and their NLSY79 matched participant-wave using a weighted Euclidean distance. Continuous covariates were z-scored by subtracting the mean and dividing by the standard deviation of the HRS sample for matching. We restricted matches to the observation with the smallest distance for each NLSY79 participant (i.e. each HRS participant can only be matched with a given NLSY79 participant once). We then selected up to ten NLSY79 participants with the smallest distance score to comprise the HRS participant’s matches.

To assure the quality of the matches between each HRS-NLSY79 pair, we considered several possible restrictions for the match quality threshold: 0.25 SD, 0.5 SD, 1.0 SD, and 1.5 SD. The wider the caliper, the larger the sample size and thus more precise the effect estimates. We interpreted the most precise estimates if the coefficients varied negligibly across matching calipers.

After employing these matching procedures for each of the six NLSY79 exposure-years, we ended up with synthetic cohort sample sizes ranging from n=46,210 NLSY79-HRS pairs for the synthetic cohort defined by the 1985 NLSY79 exposure year to n=63,091 for the 1988 exposure-year.

### Statistical Analysis

We first estimated effects within each component cohort: 1) examining the association of early adulthood through middle age alcohol use with average memory score and change in middle adulthood in NLSY79; 2) examining the association of alcohol use in middle adulthood with mid-to-late adulthood memory function and change in HRS. We estimated linear mixed effects models in both cohorts.

For analyses in NLSY79 alone, we adjusted for time-invariant confounders including age at exposure, sex, US Nativity, race/ethnicity, Armed Forces Qualification Test (measured in 1981), parental education, own education, and interview mode (in person vs not). We also adjusted for time-varying confounders using data from the wave prior to the wave the exposure was measured including religion, marital status, military service, and income (i.e. if alcohol was measured in 1983, time-varying covariates were obtained from 1982). After 2000, NLSY79 began collecting data on health and illness allowing us to further control for self-reported diabetes, self-reported hypertension, self-reported cancer, self-reported heart problems, physical activity, and general health. We controlled for a similar set of covariates in the HRS, however, covariates were measured contemporaneously with the exposure and AFQT (or any measure of earlier-life cognition) was not available.

To estimate effects of early adulthood alcohol use (as reported in NLSY79) on long-term memory change across mid-adulthood (measured in HRS), we used the synthetic cohorts of matched NLSY79/HRS pairs. We randomly assigned, without replacement, each matched pair to one of ten analytic data sets based on the matched HRS participants. If an HRS participant had fewer than ten matches of adequate quality, then the HRS participant was not included in every analytic data set. We used mixed effects models to estimate the effect of early adult alcohol use on memory level at age 65 and slope thereafter, conditional on the same set of confounding variables as those in NLSY79 with values measured from NLSY79. We then used Rubin’s rules to obtain a single estimate pooled across the ten analytic sets.^14^ We present estimates and 95% confidence intervals for coefficients pooled across mixed effects models.

Our primary parameters of interest in the synthetic cohorts were the associations of early adult alcohol use with longitudinal memory score and on memory decline across mid-to-late life, as measured by the interaction term for alcohol use and age. For comparison we also present the age slope in memory score, representing average score decline per decade of age.

## Results

Using all respondents with self-reported alcohol information, the NLSY79 sample was an average age of 22.3 (n=12,221; SD=2.3) years in 1983 and 45.2 (n=7,653; 2.2) years in 2006 (Table 1). The prevalence of current alcohol use shifted over time from 66.0% in 1983 to 51.7% in 2006. The full HRS sample at their first point of observation was 55.4 (n=21,263; 7.9) years and showed higher prevalence of alcohol use (66.3% reported use) than NLSY79 participants in 2006. Characteristics of the matched NLSY79 and HRS samples were generally similar to one another.

### Associations of early adulthood-to-midlife alcohol use with midlife memory score level and decline in NLSY79

Compared to individuals who self-reported low-to-moderate alcohol use, abstention from alcohol use at every age was neither significantly nor consistently associated with memory level in middle age among NLSY79 respondents (Table 2; Figure 1). For example, individuals who reported not drinking at ages 18-26 (1983) averaged 0.09 points (95% CI:-0.30, 0.11) worse memory in middle adulthood compared to light-to-moderate drinkers; individuals who reported not drinking at ages 41-48 (2006) averaged 0.003 points (95% CI: -0.19,0.19) better memory in middle adulthood compared to light-to-moderate drinkers. We did not observe a significant association of alcohol abstention with change in memory score in middle adulthood among NLSY79 respondents.

**Table 2:** Cohort-Specific Effects of Alcohol Use on Memory Score.

|  | Term | NLSY<br>Estimate (95%CI) for exposure round |  |  |  |  |  | HRS<br>Estimate (95%CI) |  |
| --- | --- | --- | --- | --- | --- | --- | --- | --- | --- |
|  |  | 1983<br>Age (18-26) | 1985<br>Age (20-28) | 1988<br>Age (23-31) | 1994<br>Age (29-37) | 2002<br>Age (37-45) | 2006<br>Age (41-48) | Restricting<br>to age ≤60 | All HRS<br>Follow-up* |
| Average age at alcohol self-report |  | 22 | 24 | 27 | 33 | 41 | 45 | 53 | 55 |
| Average age at cognitive assessments |  | 54 | 54 | 54 | 54 | 54 | 54 | 55 | 59 |
| Average Memory Score | No drinking | -0.093<br>(-0.297, 0.112) | 0.119<br>(-0.086, 0.324) | -0.12<br>(-0.322, 0.081) | -0.002<br>(-0.192, 0.187) | 0.1 (-0.208, 0.407) | 0.003<br>(-0.186, 0.192) | -0.107<br>(-0.223, 0.01) | -0.136<br>(-0.249, -0.023) |
|  | Heavy drinking | -0.259<br>(-0.48, -0.038) | 0.116<br>(-0.102, 0.334) | -0.15<br>(-0.345, 0.045) | -0.178<br>(-0.382, 0.026) | -0.498<br>(-0.895, -0.099) | -0.156<br>(-0.406, 0.094) | -0.264<br>(-0.446, -0.083) | -0.246<br>(-0.423, -0.069) |
| Change in Memory Score | No drinking | -0.585<br>(-1.531, 0.362) | 0.218<br>(-0.664, 1.102) | -0.826<br>(-1.683, 0.032) | -0.694<br>(-1.511, 0.124) | 0.806<br>(-1.969, 3.59) | 0.324<br>(-0.514, 1.162) | -0.093<br>(-0.4, 0.214) | -0.122<br>(-0.26, 0.016) |
|  | Heavy drinking | -1.158<br>(-2.206, -0.109) | 0.183<br>(-0.789, 1.155) | -0.846<br>(-1.68, -0.011) | -0.493<br>(-1.379, 0.392) | -1.397<br>(-5.2, 2.411) | -0.872<br>(-2.001, 0.259) | -0.454<br>(-0.946, 0.038) | -0.067<br>(-0.299, 0.164) |

Effects are relative to light-to-moderate drinkers. All models controlled for Age, Sex, US Nativity, Race/ethnicity, Parental Education, Own Education, AFQT, Religion (lagged), Marital Status (lagged), Military Service (lagged), Income (Lagged). NLSY79 models measured after 2000 and all HRS models further controlled for lagged self reported health conditions (Diabetes, Hypertension, Cancer, heart problem), lagged physical activity, and lagged general health. \*Age centered at 65

Additionally, self-reported heavy alcohol consumption at any age (except for ages 20-28) was associated with worse average memory score in middle age. NLSY79 participants who reported heavy drinking at ages 18-26 (1983) averaged 0.26 points (95% CI: -0.48, -0.04) worse memory in middle adulthood compared to light-to-moderate drinkers; individuals who reported heavy drinking at ages 41-48 (2006) averaged 0.16 points (95% CI: -0.41, 0.09) worse memory score compared to light-to-moderate drinkers. Compared to self-reported light-to-moderate drinkers, participants who reported heavy drinking exhibited a similar change in memory score in middle adulthood among NLSY79 participants.

### Associations of midlife alcohol use with mid-to-late life memory score level and decline in HRS

At their first cognitive test the mean age (IQR) of HRS participants was [55.4 (51.0,57.0)] years. Compared to light-to-moderate drinking in midlife, abstention was associated with -0.14 (95% CI: -0.25, -0.02) point lower average memory score in mid-to-late adulthood among HRS participants, and non-significantly faster rate of decline in memory score per decade of age (abstention by age interaction coefficient [-0.12 (95% CI: -0.26, 0.02)]). Heavy drinking in midlife was associated with 0.25 (95% CI: -0.42, -0.07)] point lower memory score averaged over mid-to-late life, and a non-significantly faster rate of decline in memory score per decade of age (heavy drinking by age interaction coefficient -0.07 (95% CI: -0.30, 0.16). Results were similar when restricting analyses to observations when HRS participants were age 60 or younger, to more closely mirror the age of NLSY79 participants in 2006 (Table 2; Figure 1).

### Associations of adolescent-to-midlife alcohol use with midlife-to-late life memory score in the synthetic cohorts

Covariate overlap of matching variables across NLSY79 waves compared to the HRS baseline sample are presented in Appendix Figure 1.

Self-reported abstention from alcohol use at most ages was inconsistently associated and never significantly with average memory level in mid-to-late life in the synthetic cohorts (Table 3; Figure 2). Matched-pairs who reported abstaining from alcohol at ages 18-26 averaged 0.11 points (95% CI:-0.12, 0.35) higher average memory in mid-to-late adulthood compared to light-to-moderate drinkers; matched-pairs who reported not drinking at ages 41-48 averaged 0.18 points (95% CI: -0.44,0.09) lower memory in mid-to-late adulthood compared to light-to-moderate drinkers. We did not observe a significant association of alcohol abstention with change in memory score, regardless of the age when alcohol use was reported.

**Table 3:** Associations of self-reported alcohol use with mid-to-late life cognition in synthetic cohorts.

| Term |  | NLSY exposure round in synthetic cohort with HRS outcomes |  |  |  |  |  |
| --- | --- | --- | --- | --- | --- | --- | --- |
|  |  | Estimate (95%CI) [Standard Error] |  |  |  |  |  |
|  |  | 1983 | 1985 | 1988 | 1994 | 2002 | 2006 |
| N=Unique HRS respondents |  | 6758 | 6371 | 6999 | 6783 | 6952 | 6928 |
| N=Unique NLSY respondents |  | 4683 | 4477 | 4861 | 4789 | 4838 | 4762 |
| N matched pairs |  | 54076 | 46210 | 63091 | 58349 | 62644 | 59078 |
| Average age at alcohol report |  | 22 | 24 | 27 | 33 | 41 | 45 |
| Average age at Memory Assessment |  | 59 | 59 | 59 | 59 | 59 | 59 |
| Average Memory Score | Non-drinking | 0.113<br>(-0.119,0.345) | 0.06<br>(-0.18,0.301) | -0.075<br>(-0.399,0.25) | 0.029<br>(-0.133,0.192) | -0.055<br>(-0.352,0.243) | -0.177<br>(-0.438,0.085) |
|  | Heavy drinking | 0.02<br>(-0.237,0.278) | 0.075<br>(-0.247,0.397) | -0.076<br>(-0.329,0.176) | 0.015<br>(-0.199,0.229) | 0.034<br>(-0.34,0.408) | -0.033<br>(-0.276,0.21) |
| Change in Memory | Non-drinking | 0.294<br>(-0.646,1.235) | 0.502<br>(-0.497,1.5) | -0.331<br>(-1.28,0.617) | -0.035<br>(-1.03,0.961) | -0.637<br>(-5.088,3.815) | 0.671<br>(-0.257,1.6) |
|  | Heavy drinking | -0.288<br>(-1.607,1.031) | 0.322<br>(-0.814,1.457) | -0.483<br>(-1.308,0.342) | -0.063<br>(-0.902,0.776) | -2.181<br>(-7.803,3.442) | -0.093<br>(-1.153,0.968) |

Synthetic cohorts were created for each exposure-year by matching HRS and NLSY participants based on a select set of variables suspected to confound or mediate the association between alcohol use and memory score. The set of covariates matched on may vary by exposure year; age is centered at 65. Matched pairs were quality controlled to be within 1SD of each other for each distance variable. Effects are relative to light-to-moderate drinkers.

Additionally, self-reported heavy alcohol consumption at any age was inconsistently and non-significantly associated with average memory score in middle age among matched pairs in each synthetic cohort. Matched-pairs who reported heavy drinking at ages 18-26 averaged 0.02 points (95% CI: -0.24,0.28) higher memory in middle adulthood compared to light-to-moderate drinkers; Matched-pairs who reported heavy drinking at ages 41-48 averaged 0.03 points (95% CI: -0.28,0.21) higher memory score compared to light-to-moderate drinkers. We did not observe a significant association of heavy drinking with change in memory score in middle adulthood among the matched pairs, regardless of the age of alcohol reporting.

## Discussion

In NLSY79, neither abstention from alcohol use nor heavy alcohol use were associated with midlife memory level. Conversely, in HRS both abstention and heavy drinking in midlife were associated with worse memory scores in mid-to-late-life, with heavy drinking exhibiting a stronger association. Effect estimates for midlife alcohol’s association with midlife memory score differed between the two cohorts. Patterns in average memory score were similar in the synthetic cohort with no consistent associations observed for either non-drinking or heavy drinking with average memory. Additionally, no consistent associations were found with rate of memory decline in NLSY79, HRS, nor the synthetic data, although confidence intervals were too wide to conclusively rule out a small association.

This is the first study to examine associations between prospectively reported early-adulthood alcohol use and memory score and decline in middle age and later. Our findings in the HRS are consistent with several prior studies showing that self-reported abstention from alcohol use in middle age is associated with worse cognitive outcomes.^2,5,15,16^ This result has largely been interpreted as reflecting a sick-quitter phenomenon: people who do not currently drink are assumed to be past heavy drinkers who stopped.^5,15^ However, non-drinkers are a heterogeneous group that includes former problem drinkers and those who abstain for religious or health reasons. Thus, associations of non-drinking with adverse memory may be due to an enrichment of people at higher-risk for cognitive problems, for example due to cardiovascular disease.^17–19^ In contrast, our findings in the NLSY79 showed a null association between prospectively reported abstention from alcohol decades prior to cognitive assessments, at an age when few respondents would have had major health concerns. This result may call into question simple sick-quitter explanations for the adverse associations. We cannot rule out the possibility that individuals with health concerns never initiate alcohol use.

We ran several sensitivity analyses to ensure the quality of the synthetic cohort. We tested different quality cut-offs to determine if match distance significantly affected the estimates obtained; generally, the power increased as we increased the distance cutoff from 0.25 SD to 1.5 SD with effect estimates remaining largely the same. We tried different matching sets (up to 30 matching variables) for each exposure; as we matched on more variables, the precision decreased while the estimated association remained similar. As such, for each exposure, we presented the matched set with the minimum number of matching variables that yielded an association most similar to the results obtained from a completely imputed dataset without reducing statistical power.

This study has limitations. As with all observational studies, we cannot definitely conclude that our estimated associations mirror causal effects due to the potential for unmeasured confounding. Even collected prospectively, there may be systematic error in alcohol use reporting due to desirability bias. Measures of alcohol consumption on different scales (e.g. drinks per months vs weeks) may not have a transformation that could be used to make them directly comparable.^20,21^ Inconsistencies in the magnitude of coefficients corresponding to the change in alcohol measures is difficult to interpret: this may plausibly be a measurement artifact or a true increase in the relevance of alcohol use as individuals age. Other covariates, such as own or parental education may have been affected by macro-level confounders, such as compulsory schooling laws.^22^ Though we attempted to address missing data concerns using the information we had on hand, data may not be missing at random and missingness may be affected by alcohol use or memory. Similarly, alcohol use and memory score may affect selection and survival in the sample.^23^ Validity of inferences from our synthetic cohort relies on strong assumptions about the matching algorithm. While our approach evaluated the robustness of the matching strategy to create the synthetic cohort, there is likely substantial measurement error in the matched exposure variables and there are likely to be uncontrolled biasing pathways including the potential for insufficient control for variables adjusted for in synthetic cohort models but not included in the matching process due to lack of cross-cohort availability (e.g. AFQT scores).

Key strengths were use of novel strategies and synthetic data to examine the effect of alcohol use over the life course. By using two datasets with rich data collected prospectively, we were able to examine the association of prospectively reported early adult alcohol with cognitive outcomes collected regularly in middle to late adulthood. Furthermore, while the creation of the synthetic cohort relies on several assumptions, it is reassuring that several of the different matching sets produced similar effect estimates.

Compared to low/moderate alcohol use, both alcohol abstention and heavy drinking in early to middle adulthood were not associated with worse mid-to-late life memory nor with rate of memory decline. This leaves uncertainty about the benefits of complete alcohol abstention. We find little evidence that alcohol use within the ranges reported in these cohorts substantially increased the rate of memory decline. Future studies are needed to evaluate changes in alcohol use over time and subsequent cognitive outcomes.

## Data Availability

All data produced are available online at:
https://www.bls.gov/nls/nlsy79.htm and
https://hrs.isr.umich.edu/about

https://www.bls.gov/nls/nlsy79.htm

---

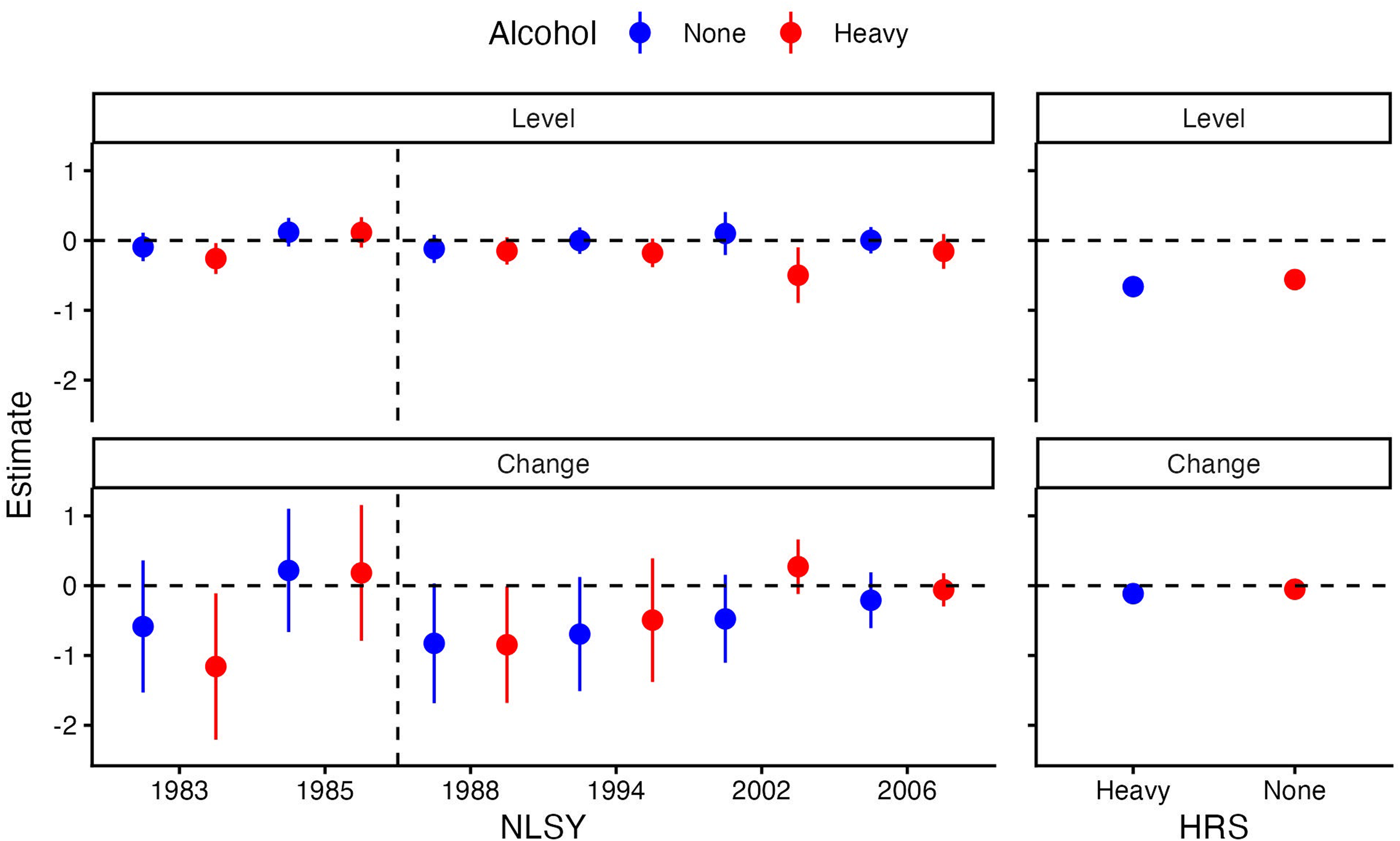

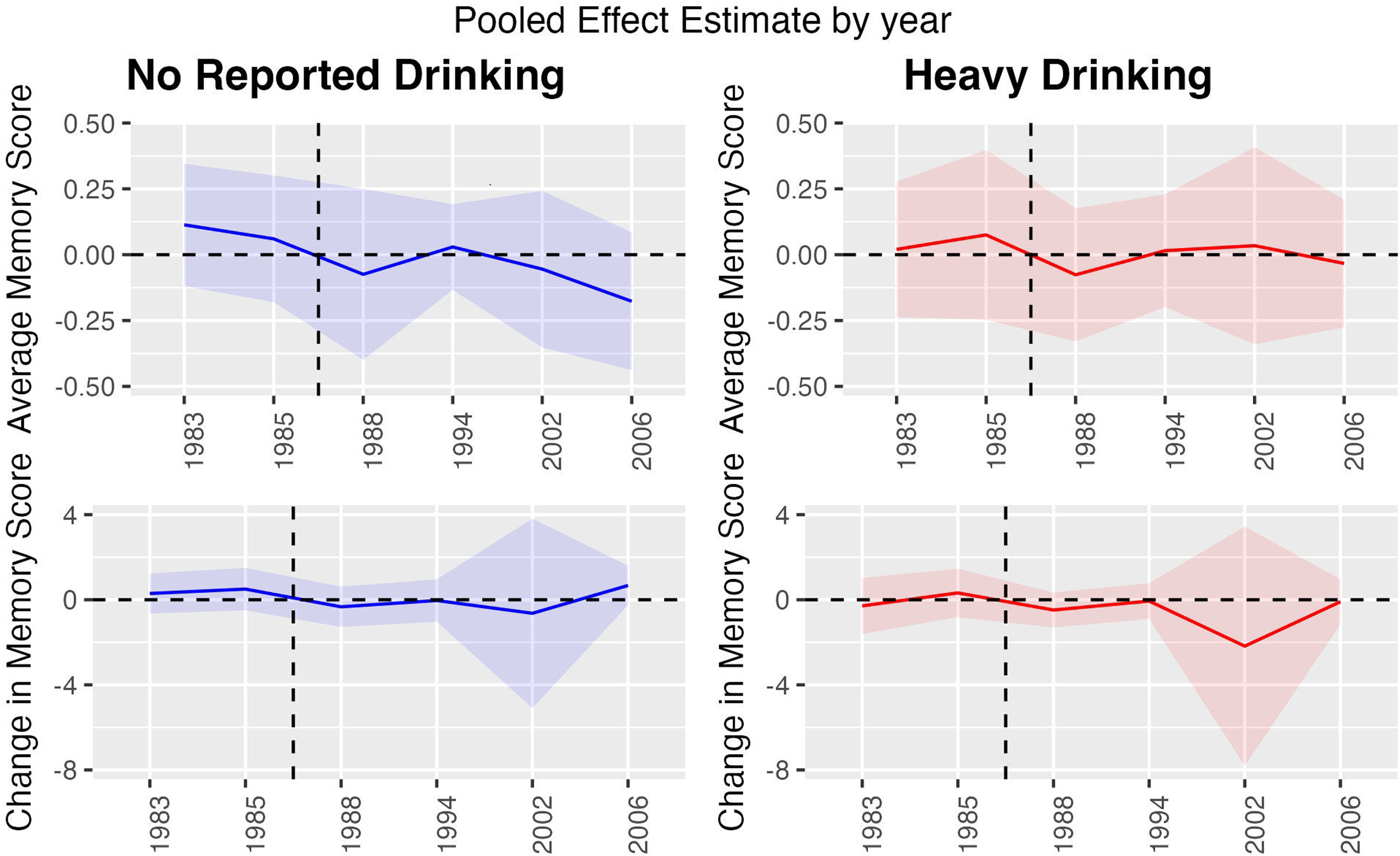

